# Epigenetic Clocks Moderate the Impact of Marital Status Transitions on Health in Older Adults

**DOI:** 10.1101/2025.06.11.25329448

**Authors:** Meng-Jung Lin

## Abstract

Chronological age is commonly used to study aging, but biological aging may more accurately reflect cumulative life experiences and psychosocial stressors. This study examines whether epigenetic clocks function as markers of resilience by assessing how marital status transitions are associated with biological aging and health outcomes in later life. Using data from 1,449 non-Hispanic White participants in the Health and Retirement Study, we analyzed thirteen epigenetic clocks derived from DNA methylation profiles. Ordinary least squares and Cox regression models assessed the associations between marital transitions, depressive symptoms, and mortality, adjusting for genetic and social factors. Interaction terms tested whether epigenetic clocks moderated these associations. Results showed that divorce and widowhood were linked to accelerated epigenetic aging. Marital status changes were associated with increased depressive symptoms but not with mortality risk. GrimAge and DunedinPACE moderated the relationship between marital disruption and depressive symptoms, while Zhang and GrimAge moderated the relationship with mortality risk. Biologically older individuals, particularly men, exhibited greater resilience to these transitions. These findings raise the possibility that epigenetic clocks reflect accumulated life experiences and psychosocial adaptation, potentially including elements of resilience in older adults.

## Introduction

Aging has long fascinated human societies, from Aristotle’s philosophical inquiries into the nature of biological deterioration to Emperor Qin Shi Huang’s legendary quest for immortality. Across time and cultures, the collective aspiration has not simply been to halt chronological aging but to extend vitality and preserve youthfulness. Although chronological age is conventionally treated as a neutral administrative marker, it carries profound biological, psychological, and social meanings (1–3). Externally, age structures legal entitlements, governs transitions into and out of institutional roles, and shapes interpersonal expectations. Internally, individuals construct their identities in part through the lens of age. While positive stereotypes emphasize wisdom, reliability, and emotional maturity, negative perceptions focus on rigidity, cognitive decline, and physical frailty (4). These stereotypes not only shape how older adults are treated by others but may also be associated with self-perceptions, with potential implications for health, social integration, and psychological well-being. Recognizing the positive aspects of aging, particularly the accumulation of resilience and adaptive skills, provides a fuller and more balanced understanding of aging.

One key positive trait associated with aging is resilience, broadly defined as the capacity to adapt successfully to adversity (5). Older adults often draw upon a reservoir of life experiences, coping strategies, and emotional regulation skills that younger individuals have had less opportunity to cultivate (6,7). Life experiences, both positive and negative, contribute to a form of psychological maturity that supports adaptive functioning under conditions of stress. Consistent with the adage “what doesn’t kill you makes you stronger,” many older adults demonstrate an enhanced ability to navigate personal, relational, and health-related adversities. Understanding resilience as an outcome of accumulated life experience reframes aging not as an inevitable decline but as a process of growth and adaptation.

Building on this conceptualization, several theoretical frameworks help to contextualize resilience within aging. Resiliency Theory (8) emphasizes the importance of internal and external protective factors, such as coping skills, competence, and supportive relationships, that enable individuals to thrive despite adversity. Psychological resilience has also been conceptualized as fundamentally tied to emotion regulation, with affect regulation playing a central role in resilient outcomes (9). Extending these perspectives into gerontology, (10) provided a conceptual analysis of resilience in aging, emphasizing how acceptance, adaptation, and enriched coping strategies contribute to successful aging. Collectively, these frameworks underscore that aging is not merely a period of decline but also one of psychological growth, where older adults draw on a lifetime of emotional and social resources to sustain well-being amid life’s challenges.

Viewing aging as a multidimensional process offers a more integrated perspective on how individuals adapt to age-related challenges. Recent theoretical frameworks emphasize the biological, psychological, and social dimensions of aging (2,4). Biological age reflects the physiological condition of the body, psychological age refers to self-perceptions of aging, and social age encompasses culturally constructed roles and expectations tied to chronological milestones. These dimensions interact in complex ways. For example, an individual may have a younger biological age relative to peers but feel socially marginalized due to age-related stereotypes. Conversely, someone with significant health issues may nonetheless maintain a strong sense of psychological youth and active social engagement.

A significant development in the biological measurement of aging has been the introduction of epigenetic clocks. These biomarkers estimate biological age by analyzing patterns of DNA methylation across the genome (11). Although initially conceptualized as purely biological indicators, a growing body of research suggests that epigenetic clocks are sensitive to social environments and life course exposures. Socioeconomic disadvantage, marital status, occupational stress, lifestyle behaviors, and psychosocial adversity have all been associated with differences in epigenetic aging (12–15). For instance, using data from the National Longitudinal Study of Adolescent to Adult Health (Add Health), (16) showed that educational attainment, income, health behaviors, and stress exposure robustly predict variation in epigenetic age acceleration across multiple clocks. Their findings further highlight the extent to which these biomarkers reflect both biological and social processes accumulated over the life course.

The plasticity of biological age aligns closely with life course theories that emphasize cumulative exposure, adaptation, and individual agency (17,18). Biological systems respond dynamically to social and environmental contexts, adjusting to both adversity and support over time. Within this framework, major life transitions such as changes in marital status represent critical junctures that can recalibrate health trajectories both psychologically and biologically. Marital transitions, in particular, have been extensively examined in relation to health and longevity. Marriage is associated with better mental health, reduced risk of chronic disease, and longer survival, especially for men (19,20). In contrast, divorce and widowhood are often linked to increased psychological distress, diminished social and financial resources, and elevated mortality risk (21–26). These transitions are not merely psychosocial events; they also leave biological imprints. Studies have found that spousal loss is associated with physiological dysregulation, increased inflammation, and markers of accelerated aging (27–29).

Although some research has begun to examine associations between marital status and epigenetic aging, fewer studies have focused on whether specific life transitions such as divorce and widowhood contribute to biological age acceleration when viewed through a life course lens. To establish a baseline association and motivate subsequent analyses of resilience, this study begins by testing the following hypothesis:

H1: Individuals who experience marital dissolution (divorce or widowhood) exhibit greater biological age acceleration compared to those who remain partnered.

Psychological resilience has emerged as a key factor in moderating the biological consequences of life stress. Recent empirical research has expanded this understanding by exploring how resilience processes intersect with biological aging. Cumulative stress exposures, including trauma, discrimination, and chronic adversity, have been consistently linked to accelerated epigenetic aging (30,31). However, psychological resilience factors, particularly emotion regulation and self-control, can buffer these associations. Individuals with stronger emotion regulation or higher self-control exhibit slower DNA methylation age acceleration, suggesting that psychosocial resources serve as biological buffers (30). A study using data from the Health and Retirement Study found that individuals with higher psychological resilience scores experienced slower epigenetic aging across multiple clock measures (32). These findings underscore the plasticity of biological aging and point to resilience as a promising target for interventions aimed at promoting healthier aging trajectories (33–35).

Although aging is often portrayed as a period of vulnerability, a growing body of research highlights the emergence of psychological strengths across the life course. Emotional regulation, for example, tends to improve with age, as older adults become more adept at sustaining positive affect and downregulating negative emotions (36,37). This capacity plays a critical role in buffering the effects of stress on health (38). Building on these findings, researchers have increasingly framed resilience as a psychological resource that slows biological aging (30,32). However, this preventive framing may not fully capture the complexity of aging in later life. In some cases, an older epigenetic age may not solely indicate vulnerability. It may also reflect the cumulative imprint of life experiences and the adaptive strategies developed to manage them. From this perspective, epigenetic clocks could serve not only as intervention targets but also as indicators of resilience shaped by lived experience. Resilience, in this broader view, encompasses both psychological and biological dimensions. For instance, studies of individuals exposed to early life adversity, such as combat veterans, suggest that hardship can foster developmental maturity and greater resilience in later life (39).

Similarly, research by (6) shows that older adults often draw on a repertoire of coping strategies accumulated over the life course to manage new challenges. This perspective suggests that biological aging, as measured by epigenetic clocks, may capture not only cumulative burden but also accumulated resilience. If epigenetic clocks reflect both deterioration and adaptation, they may moderate the relationship between marital status change and later health outcomes. This leads to the second hypothesis:

H2: Epigenetically older individuals exhibit greater resilience to marital status change compared to their epigenetically younger counterparts.

Gender differences are likely to structure these processes. Men and women differ in both the experience and consequences of marital transitions. Men are more vulnerable to social isolation and health decline following marital loss, whereas women, despite often facing greater economic hardship, may benefit from stronger social networks and emotional coping resources (19,23,40). Widows may reframe solitude as autonomy, while widowers are more likely to experience it as deprivation (40). These gendered patterns suggest that the buffering effects of resilience, as indexed by epigenetic clocks, may operate differently by gender. Accordingly, the third hypothesis is proposed:

H3: The moderating effects of epigenetic clocks on mental health and mortality following marital status change are stronger among men than among women.

Using data from the Health and Retirement Study, this study examines how marital status transitions are associated with biological aging and whether epigenetic clocks, conceptualized as proxies for accumulated life experiences, moderate the impact of these transitions on depressive symptoms and mortality. In doing so, it explores whether epigenetic clocks function not only as indicators of aging but also as potential markers of accumulated psychosocial adaptation. By integrating biological, psychological, and social frameworks, the analysis contributes to a more nuanced understanding of resilience in later life and highlights the complex, multidimensional nature of aging.

## Methods

### Data source

Data for this study were drawn from the Health and Retirement Study (HRS) (https://hrs.isr.umich.edu/), a longitudinal, nationally representative survey initiated in 1992 by the Institute for Social Research (ISR) at the University of Michigan and sponsored by the National Institute on Aging (NIA) (41). The HRS tracks the lives of U.S. adults aged 50 and older through biennial interviews, with the primary objective of capturing the social, economic, and health-related factors contributing to retirement and aging. The de-identified data used in this study were accessed in December 2023; at no point did we have access to any information that could identify individual participants.

In addition to survey data, DNA methylation data were collected from a subsample of HRS participants (N = 4,104) who took part in the 2016 Venous Blood Study (42). During scheduled home visits, trained phlebotomists collected six tubes of blood, including a 10 mL EDTA whole blood tube. The EDTA tubes were shipped to the University of Minnesota laboratory for processing and DNA extraction. DNA methylation was assayed using the Infinium MethylationEPIC BeadChip, resulting in high-quality data for 4,018 participants after quality control procedures.

To incorporate genetic factors as control variables, the study also utilized genetic data from the HRS (43). Saliva samples were collected from participants during the 2006, 2008, and 2012 survey waves and genotyped using the Illumina HumanOmni2.5 BeadChips at the Center for Inherited Disease Research. Of the more than 19,000 collected samples, approximately 15,000 passed quality control checks conducted at the Genetics Coordinating Center at the University of Washington.

Additionally, the HRS includes the RAND HRS Family Data (44), which provides harmonized information on respondents’ children, parents, and siblings across multiple waves. This information enabled the examination of how social support related to health outcomes through measures such as geographic proximity and financial transfers between children and parents, enhancing the analysis of health outcomes.

By merging DNA methylation data, GWAS data, and detailed survey information, this study integrates epigenetic, genetic, and social measures to test the three research hypotheses.

### Measures

The study considered the following measures:

### Epigenetic clocks

Previous research has identified genomic regions where methylation changes correlate with chronological age or, more recently, health outcomes associated with aging (42). Thirteen epigenetic clocks were constructed by the HRS, including nine first-generation clocks trained on chronological age and four second-generation clocks trained on health-related outcomes (13).

These clocks include Horvath 1, Hannum, Lin, Weidner, Vidal-Bralo, Horvath 2, EpiTOC (Yang), Bocklandt, Zhang, Levine-PhenoAge, Lu-GrimAge, and DunedinPACE. DNAm age was calculated as a weighted sum of methylation values at age-associated CpG sites, with weights derived through machine learning models. Table 1 provides details for the thirteen epigenetic clocks. These clocks served as dependent variables in the first set of analyses and as independent variables and moderators in the second set of analyses predicting health outcomes.

**Table 1.** Details of The Thirteen Epigenetic Clocks in HRS.

| Clock name | First author (year) | Training phenotype | Tissue type(s) | Number of CpG sites | Unit of measurement | Range in HRS |
| --- | --- | --- | --- | --- | --- | --- |
| **Horvath 1** | Horvath (2013) | Chronological age | 51 tissues/cells | 353 | Years | 23.3 – 114.5 |
| **Hannum** | Hannum (2013) | Chronological age | Whole blood | 71 | Years | 25.1 – 107.8 |
| **Levine** | Levine (2018) | Phenotypic age | Whole blood | 513 | Years | 26.7 – 101.7 |
| **Horvath 2** | Horvath (2018) | Chronological age | Skin | 391 | Years | 37.0 – 101.3 |
| **Lin** | Lin (2015) | Chronological age | Whole blood | 99 | Years | 1.9 – 133.3 |
| **Weidner** | Weidner (2014) | Chronological age | Whole blood | 3 | Years | 25.2 – 148.9 |
| **VidalBralo** | Vidal-Bralo (2016) | Chronological age | Whole blood | 8 | Years | 36.5 – 109.9 |
| **EpiTOC** | Yang (2016) | Chronological age | Whole blood | 385 | Average % DNAm | 0.0 – 0.2 |
| **Zhang** | Zhang (2017) | Mortality risk (time-to-death) | Whole blood | 10 | A unit of mortality risk score | -2.5 – 0.6 |
| **Bocklandt** | Bocklandt (2011) | Chronological age | Saliva | 2 | Average % DNAm | 0.1 – 0.9 |
| **Garagnani** | Garagnani (2012) | Chronological age | Whole blood | 1 | Average % DNAm | 0.4 – 1.0 |
| **GrimAge** | Lu (2019) | Mortality risk (time-to-death) | 7 plasma proteins, smoking pack years | 1030 | Years | 42.7 – 99.6 |
| **DunedinPACE** | Belsky (2020) | Rate of change across 18 biomarkers | Whole blood | 46 | Rate of aging in years | 0.7 – 1.5 |

Sources: Crimmins et al. (2020), Schmitz et al. (2022)

### Dependent variables

Depressive Symptoms: The CESD (Center for Epidemiologic Studies Depression Scale) score was used to assess depressive symptoms. An abridged eight-item version of the CESD was used in HRS, asking respondents whether, during the past week, they felt depressed, felt that everything they did was an effort, experienced restless sleep, felt unable to get going, felt lonely, enjoyed life, felt sad, and were happy. Responses were coded as “yes” or “no.” Positive items were reverse-coded, and the items were summed to create the CESD score, with higher scores indicating more depressive symptoms. The second set of analyses used the 2020 wave CESD score as the outcome.

Mortality: HRS documented respondents’ age at death in months. Death dates were collected from Exit Interviews or based on spouse-reported information. The age-at-death variable was calculated by RAND using birth and death dates, measured in months. Because the measurement of epigenetic clocks is essential for testing the study hypotheses, the analytic sample was restricted to individuals with methylation data, with baseline set at their age in months at the beginning of the 2016 interview wave. Observations were right-censored at the end of the 2020 wave.

### Independent variables

Marital Status: Respondents reported their current marital status as married, married with spouse absent, partnered, separated, divorced, separated/divorced, widowed, or never married. To avoid sparse categories and to facilitate interpretation, marital status was recoded into five categories: married, partnered, separated/divorced, widowed, and never married. Marital status measured in 2014 was used to predict epigenetic aging, while marital status in 2016 was used to predict later outcomes.

Marital Status Change: Marital status change was conceptualized as a life event to examine resilience in later life. Since epigenetic clocks were measured in 2016, marital status change was assessed by comparing marital status across 2016, 2018, and 2020. For mortality analyses, marital status change was based on comparisons across 2012, 2014, and 2016, given that death could occur between 2016 and 2020. A change in marital status was coded as 1, while no change was coded as 0.

### Control variables

Control variables included polygenic scores (PGS) for longevity, number of children ever born, age at first birth, and depressive symptoms, as well as measures of social support, number of children, health behaviors, educational attainment, highest parental education, total household wealth, retirement status, chronological age, biological sex, family size, number of living siblings, cohort membership, religious affiliation, and population stratification.

A polygenic score (PGS), also known as a polygenic risk score (PRS), summarizes an individual’s genetic predisposition to a particular trait or disease, based on the cumulative effect of multiple genetic variants. It is calculated as the weighted sum of risk alleles: 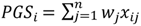 where 𝑖 indexes individuals, 𝑗 indexes SNPs, 𝑤 denotes the weight for each SNP, and 𝑥 is the number of risk alleles. Polygenic scores were constructed by the HRS (43) using summary statistics from GWAS studies on longevity (45), reproductive behaviors (46), and depressive symptoms (47). All PGS were standardized to ease interpretation.

The longevity PGS was included to control for genetic predispositions to longer life, which may confound associations between epigenetic age and mortality. PGS for number of children ever born and age at first birth were included to account for potential marriage selection processes, particularly given generational patterns linking fertility and marriage. Depressive symptoms PGS was controlled because it may be related to both the biological and psychological outcomes of interest. Including these genetic controls helped isolate the contribution of environmental and social experiences to observed patterns in epigenetic aging.

Population Stratification: To control for ancestral differences in allele frequencies that could bias genetic associations, the first ten principal components (PCs) from a principal component analysis of genetic ancestry were included in all regression models, as recommended by (48).

Social Support and Health Behaviors: Social support was measured using three variables: the number of children living within 10 miles, the dollar amount of financial transfers received from children, and the dollar amount of financial transfers given to children. Health behaviors were captured using measures of vigorous physical activity, alcohol consumption, and smoking history. Vigorous physical activity was assessed with the question: “How often do you take part in sports or activities that are vigorous, such as running, swimming, cycling, aerobics or gym workouts, tennis, or digging with a spade or shovel?” Responses ranged from “every day” to “never.” For analysis, responses were dichotomized into frequent (every day or more than once per week = 1) and infrequent (all other responses = 0). Alcohol consumption was assessed by the question “Do you ever drink any alcoholic beverages, such as beer, wine, or liquor?” (coded yes = 1, no = 0). Smoking history was assessed by the question “Have you ever smoked cigarettes?” (coded yes = 1, no = 0).

Cohort Membership: Respondents were grouped into three birth cohorts: older (AHEAD <1924, CODA 1924–1930, HRS 1931–1941), middle (WB 1942–1947), and younger (EBB 1948–1953, MBB 1954–1959, LBB 1960–1965).

### Analytical strategies

Two sets of analyses were conducted to test the study hypotheses. First, ordinary least squares (OLS) regression models were used to examine the effect of marital status on epigenetic aging. Social factors and polygenic scores were included as covariates to account for potential selection effects.

Second, epigenetic clocks were treated as markers of accumulated life experiences and were used to predict depressive symptoms, expected mortality, and mortality. OLS models were employed to predict depressive symptoms, while Cox proportional hazards models were used to predict mortality risk. In addition to assessing the main effects of epigenetic clocks on health outcomes, interaction terms between epigenetic clocks and marital status change were included to test moderation hypotheses. Significant interaction effects would suggest that epigenetic clocks may function as a resilience-related resource, moderating the impact of marital status changes on depressive symptoms and mortality.

To assess gender differences as proposed in Hypothesis 3, all analyses were stratified by biological sex, with separate models estimated for men and women.

Finally, because both epigenetic clocks and polygenic scores were developed primarily in European ancestry populations, analyses were restricted to non-Hispanic White respondents to minimize bias from population stratification. Starting with 4,018 respondents who had DNA-methylation data, 2,314 met the ancestry criterion and had valid polygenic scores. Additional exclusions for missing CESD data in 2020 (494), incomplete marital-status history between 2014 and 2020 (64), missing measures of social support (215), missing health-behavior data (16), and missing demographic covariates (76) yielded a final analytic sample of 1,449 for the models predicting epigenetic aging and depressive symptoms. The mortality analysis began with the same 4,018 respondents. After removing 330 individuals who lacked duration-to-death or censoring information, 2,158 remained. Subsequent exclusions for missing polygenic scores (1,530), marital-status history (23), social support variables (262), health-behavior data (17), and demographic covariates (99) produced a final sample of 1,757 for the Cox proportional hazards models.

We used ChatGPT (OpenAI, GPT-4o) to improve clarity and grammar. All content was reviewed and finalized by the authors.

## Results

### Descriptive statistics

Table 2 presents descriptive statistics for the variables used in the CESD score analysis. On average, the epigenetic ages of the sample range from approximately 55 to 68 years, slightly younger than the respondents’ mean chronological age of 70.87 years. Women generally exhibited younger epigenetic ages compared to men. The mean CESD score in 2020 was 1.30, with women reporting higher CESD scores than men in both 2016 and 2020. In total, 12.9% of the analytic sample died between the 2016 and 2020 interview waves, with a slightly higher mortality rate among men (14.3%) compared to women (11.9%).

**Table 2.** Descriptive Statistics of the Variables (Frequency (%) or Mean (SD)) (HRS)

|  | All | Male | Female |
| --- | --- | --- | --- |
| **N** | **1,449 (100.0%)** | **598 (41.3%)** | **851 (58.7%)** |
| Epigenetic Clocks |  |  |  |
| Horvath 1 | 67.021 (8.839) | 68.078 (8.629) | 66.278 (8.914) |
| Hannum | 55.745 (8.570) | 57.247 (8.431) | 54.689 (8.514) |
| Levine | 58.070 (9.335) | 59.345 (8.700) | 57.175 (9.662) |
| Horvath 2 | 70.806 (8.168) | 71.778 (8.115) | 70.123 (8.140) |
| Lin | 59.761 (10.494) | 60.855 (10.236) | 58.993 (10.611) |
| Weidner | 67.295 (11.343) | 68.050 (11.160) | 66.764 (11.446) |
| Vidal-Bralo | 64.361 (5.878) | 65.664 (5.621) | 63.446 (5.886) |
| EpiTOC (Yang) | 0.067 (0.017) | 0.066 (0.018) | 0.067 (0.017) |
| Zhang | -1.088 (0.440) | -0.954 (0.403) | -1.183 (0.441) |
| Bocklandt | 0.381 (0.072) | 0.367 (0.074) | 0.391 (0.069) |
| Garagnani | 0.723 (0.070) | 0.722 (0.070) | 0.724 (0.069) |
| GrimAge | 68.404 (8.040) | 70.787 (7.653) | 66.730 (7.886) |
| DunedinPACE | 1.063 (0.089) | 1.077 (0.089) | 1.054 (0.087) |
| CESD Score |  |  |  |
| CESD Score in 2020 | 1.302 (1.925) | 1.074 (1.798) | 1.462 (1.994) |
| CESD Score in 2016 | 1.046 (1.726) | 0.789 (1.444) | 1.227 (1.880) |
| Death (Total N=1,757; M: N=743; F: N=1,014) |  |  |  |
| No | 1,530 (87.1%) | 637 (85.7%) | 893 (88.1%) |
| Yes | 227 (12.9%) | 106 (14.3%) | 121 (11.9%) |
| Duration | 45.445 (8.613) | 44.603 (9.511) | 46.062 (7.839) |
| Marital Status Change (2016, 18, 20) |  |  |  |
| No | 1,298 (89.6%) | 538 (90.0%) | 760 (89.3%) |
| Yes | 151 (10.4%) | 60 (10.0%) | 91 (10.7%) |
| Marital Status |  |  |  |
| Marital Status in 2020 |  |  |  |
| Married | 905 (62.5%) | 450 (75.3%) | 455 (53.5%) |
| Partnered | 56 (3.9%) | 28 (4.7%) | 28 (3.3%) |
| Separated/Divorced | 145 (10.0%) | 51 (8.5%) | 94 (11.0%) |
| Widowed | 335 (23.1%) | 67 (11.2%) | 268 (31.5%) |
| Never Married | 8 (0.6%) | 2 (0.3%) | 6 (0.7%) |
| Marital Status in 2018 |  |  |  |
| Married | 963 (66.5%) | 475 (79.4%) | 488 (57.3%) |
| Partnered | 53 (3.7%) | 26 (4.3%) | 27 (3.2%) |
| Separated/Divorced | 150 (10.4%) | 50 (8.4%) | 100 (11.8%) |
| Widowed | 275 (19.0%) | 46 (7.7%) | 229 (26.9%) |
| Never Married | 8 (0.6%) | 1 (0.2%) | 7 (0.8%) |
| Marital Status in 2016 |  |  |  |
| Married | 1,000 (69.0%) | 487 (81.4%) | 513 (60.3%) |
| Partnered | 50 (3.5%) | 29 (4.8%) | 21 (2.5%) |
| Separated/Divorced | 155 (10.7%) | 49 (8.2%) | 106 (12.5%) |
| Widowed | 236 (16.3%) | 32 (5.4%) | 204 (24.0%) |
| Never Married | 8 (0.6%) | 1 (0.2%) | 7 (0.8%) |
| Marital Status in 2014 |  |  |  |
| Married | 1,038 (71.6%) | 501 (83.8%) | 537 (63.1%) |
| Partnered | 47 (3.2%) | 26 (4.3%) | 21 (2.5%) |
| Separated/Divorced | 154 (10.6%) | 52 (8.7%) | 102 (12.0%) |
| Widowed | 201 (13.9%) | 18 (3.0%) | 183 (21.5%) |
| Never Married | 9 (0.6%) | 1 (0.2%) | 8 (0.9%) |
| Number of Children Ever Born |  |  |  |
| 0 | 87 (6.0%) | 40 (6.7%) | 47 (5.5%) |
| 1 | 192 (13.3%) | 81 (13.5%) | 111 (13.0%) |
| 2 | 552 (38.1%) | 223 (37.3%) | 329 (38.7%) |
| 3 | 350 (24.2%) | 134 (22.4%) | 216 (25.4%) |
| 4 and more | 268 (18.5%) | 120 (20.1%) | 148 (17.4%) |
| Health Lifestyles |  |  |  |
| Vigorous Physical Activity (2016) |  |  |  |
| No | 1,036 (71.5%) | 398 (66.6%) | 638 (75.0%) |
| Yes | 413 (28.5%) | 200 (33.4%) | 213 (25.0%) |
| Ever Drinks Any Alcohol (2016) |  |  |  |
| No | 557 (38.4%) | 219 (36.6%) | 338 (39.7%) |
| Yes | 892 (61.6%) | 379 (63.4%) | 513 (60.3%) |
| Ever Smokes (2016) |  |  |  |
| No | 656 (45.3%) | 213 (35.6%) | 443 (52.1%) |
| Yes | 793 (54.7%) | 385 (64.4%) | 408 (47.9%) |
| Vigorous Physical Activity (2014) |  |  |  |
| No | 1,008 (69.6%) | 381 (63.7%) | 627 (73.7%) |
| Yes | 441 (30.4%) | 217 (36.3%) | 224 (26.3%) |
| Ever Drinks Any Alcohol (2014) |  |  |  |
| No | 538 (37.1%) | 206 (34.4%) | 332 (39.0%) |
| Yes | 911 (62.9%) | 392 (65.6%) | 519 (61.0%) |
| Ever Smokes (2014) |  |  |  |
| No | 656 (45.3%) | 213 (35.6%) | 443 (52.1%) |
| Yes | 793 (54.7%) | 385 (64.4%) | 408 (47.9%) |
| Polygenic Score |  |  |  |
| Longevity PGS | 0.020 (0.972) | 0.021 (0.997) | 0.020 (0.955) |
| Depressive Symptom PGS | -0.042 (1.013) | -0.056 (0.977) | -0.031 (1.039) |
| Number of Children Ever Born PGS | 0.017 (0.974) | 0.033 (0.994) | 0.006 (0.960) |
| Age at First Birth PGS | 0.049 (0.976) | 0.050 (0.983) | 0.048 (0.972) |
| Social Support (Only available in 2014) |  |  |  |
| Number of Children Living within 10 Miles | 0.798 (1.005) | 0.799 (1.064) | 0.797 (0.962) |
| Amount of Transfer from Children | 0.003 (0.068) | 0.005 (0.104) | 0.001 (0.011) |
| Amount of Transfer to Children | 0.060 (0.241) | 0.081 (0.333) | 0.045 (0.142) |
| Socioeconomic Background |  |  |  |
| Years of Education | 13.761 (2.266) | 13.915 (2.395) | 13.653 (2.165) |
| Parental Education | 11.977 (2.985) | 12.100 (2.982) | 11.891 (2.986) |
| Total Wealth |  |  |  |
| Total Wealth (2016) | 7.138 (13.293) | 7.558 (13.238) | 6.842 (13.330) |
| Total Wealth (2014) | 6.630 (12.981) | 7.104 (14.027) | 6.298 (12.191) |
| Retirement Status |  |  |  |
| Retirement Status (2016) |  |  |  |
| Not Retired | 320 (22.1%) | 139 (23.2%) | 181 (21.3%) |
| Completely Retired | 883 (60.9%) | 341 (57.0%) | 542 (63.7%) |
| Partly Retired | 219 (15.1%) | 118 (19.7%) | 101 (11.9%) |
| Question Irrelevant | 27 (1.9%) | 0 (0.0%) | 27 (3.2%) |
| Retirement Status (2014) |  |  |  |
| Not Retired | 407 (28.1%) | 165 (27.6%) | 242 (28.4%) |
| Completely Retired | 741 (51.1%) | 292 (48.8%) | 449 (52.8%) |
| Partly Retired | 266 (18.4%) | 139 (23.2%) | 127 (14.9%) |
| Question Irrelevant | 35 (2.4%) | 2 (0.3%) | 33 (3.9%) |
| Demographic Characteristics |  |  |  |
| Age |  |  |  |
| Age at 2016 | 70.868 (8.788) | 71.263 (8.372) | 70.591 (9.063) |
| Age at 2014 | 68.697 (8.812) | 69.092 (8.392) | 68.420 (9.090) |
| Cohort |  |  |  |
| Old | 574 (39.6%) | 251 (42.0%) | 323 (38.0%) |
| Middle | 298 (20.6%) | 124 (20.7%) | 174 (20.4%) |
| Young | 577 (39.8%) | 223 (37.3%) | 354 (41.6%) |
| Family Size |  |  |  |
| Family Size (2016) | 2.073 (0.873) | 2.144 (0.748) | 2.024 (0.949) |
| Family Size (2014) | 2.115 (0.907) | 2.179 (0.727) | 2.071 (1.013) |
| Number of Living Siblings |  |  |  |
| Number of Living Siblings (2016) | 2.411 (1.890) | 2.351 (1.840) | 2.452 (1.925) |
| Number of Living Siblings (2014) | 2.427 (1.904) | 2.363 (1.861) | 2.471 (1.934) |
| Religious Affiliation |  |  |  |
| Protestant | 932 (64.3%) | 371 (62.0%) | 561 (65.9%) |
| Catholic | 337 (23.3%) | 132 (22.1%) | 205 (24.1%) |
| None or No Preference | 148 (10.2%) | 80 (13.4%) | 68 (8.0%) |
| Other | 32 (2.2%) | 15 (2.5%) | 17 (2.0%) |

Regarding marital status, about 70% of respondents reported being married in 2016, a figure that declined to 62% by 2020, primarily due to an increase in widowhood. Approximately 10% of respondents experienced a change in marital status between 2016 and 2020. In 2020, around 4% of respondents were partnered, 10% were separated or divorced, 20% were widowed, and fewer than 1% had never married. Given the rising prevalence of widowhood among older adults, understanding the potential health and mental well-being consequences of losing a spouse is critical. Accordingly, the subsequent analyses examine how marital status is related to biological aging and how individuals’ life experiences, as measured by epigenetic clocks, may help buffer the impact of this stressful transition.

### Marital status, polygenic scores, other social factors, and epigenetic aging

Table 3 presents the models examining the associations between marital status and epigenetic aging. Overall, marital status showed limited associations with epigenetic clocks after controlling for polygenic scores and social factors. However, for two second-generation clocks, GrimAge and DunedinPACE, which are known to be more responsive to sociodemographic exposures (13), widowhood was associated with significantly older epigenetic ages compared to being married.

**Table 3.**
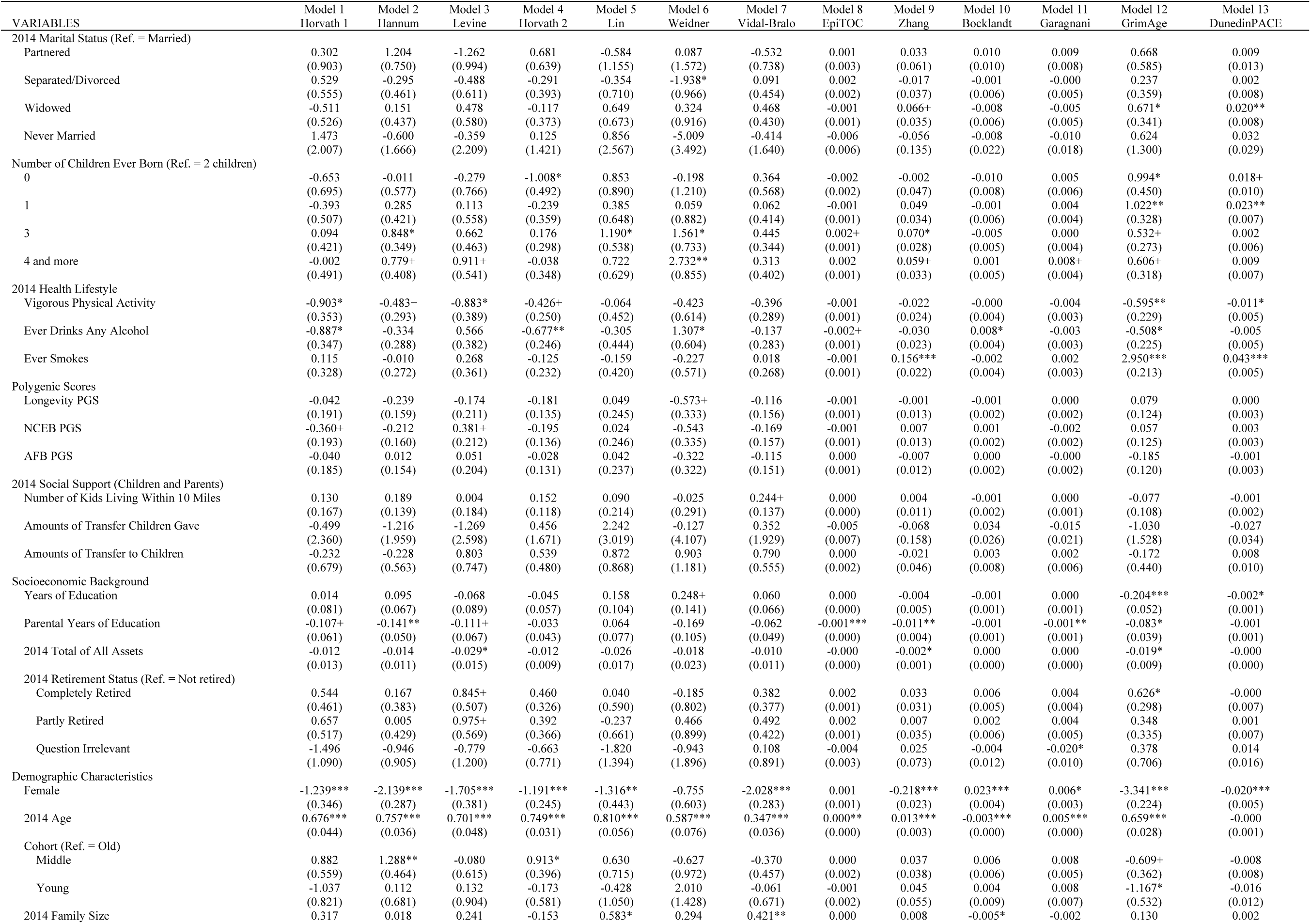
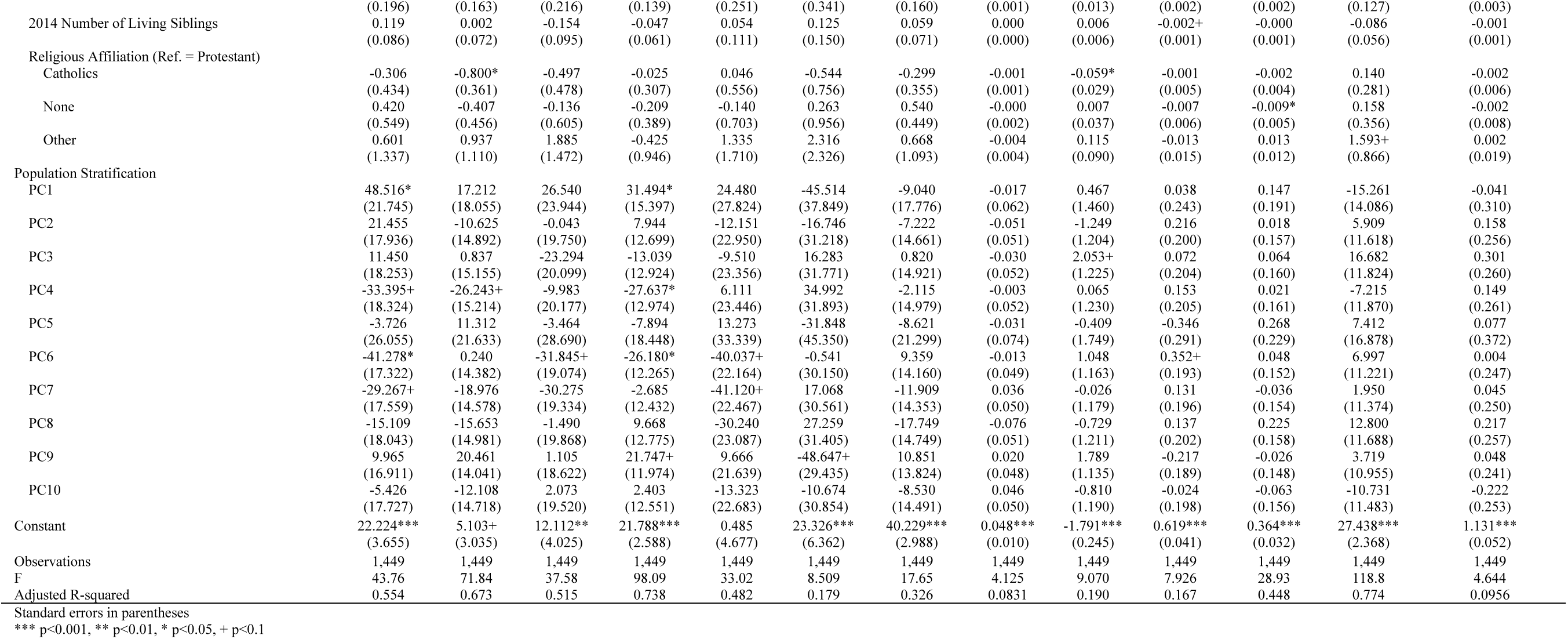
Ordinary Least Squares Models Using Marital Status, PGSs, and Social Factors to Predict Epigenetic Clocks (HRS)

In gender-stratified models (see S2 Table), widowed women exhibited significantly older DunedinPACE clocks, while separated or divorced women had elevated EpiTOC (Yang) clocks compared to their married counterparts. Among men, widowers demonstrated significantly older Levine and Vidal-Bralo clocks relative to married men. Notably, although the coefficient for being widowed was negative in the model predicting the Bocklandt clock, prior research indicates that this clock tends to be negatively correlated with other clocks (13). Divorce was not significantly associated with epigenetic aging among men; in fact, the coefficients for being divorced were negative across several models.

Beyond marital status, several social factors were significantly related to biological aging. Having three children was associated with increased biological age, particularly among women. Health behaviors followed expected patterns: frequent vigorous physical activity was associated with slower epigenetic aging, while smoking was linked to accelerated aging. Having ever consumed alcohol showed inconsistent associations with the clocks, which may reflect that the measure captured lifetime experimentation rather than drinking intensity or frequency. Occasional alcohol use may be linked to broader social engagement and openness to experience, while heavier drinking likely contributes to biological aging.

Parental education emerged as a consistent protective factor across clocks, with higher parental education associated with younger biological age. Demographically, women exhibited younger biological ages compared to men across most clocks. Chronological age was positively associated with most epigenetic clocks, although it was negatively associated with the Bocklandt clock and unrelated to DunedinPACE, consistent with the latter’s focus on accelerated biological aging rather than chronological age per se.

Regarding genetic factors, polygenic scores for longevity, number of children ever born, and age at first birth were not significantly associated with biological aging. In summary, after accounting for genetic predispositions, social factors such as marital status, health behaviors, and socioeconomic background played important roles in shaping biological aging. These findings provide empirical support for Hypothesis 1.

### Epigenetic aging, marital status change, and CESD score

S3 Table presents models examining the main effects of epigenetic clocks and marital status change on CESD scores. Overall, with the exception of GrimAge and DunedinPACE, the other eleven clocks were not significantly associated with depressive symptoms. Both GrimAge and DunedinPACE showed positive associations, indicating that accelerated epigenetic aging corresponded with higher CESD scores. In contrast, marital status change was consistently associated with an increase of approximately 0.52 points in CESD scores across models, suggesting that experiencing a marital transition may be linked to poorer mental health in later life.

Gender-stratified analyses are shown in S5 Table. Among men, none of the epigenetic clocks exhibited significant associations with CESD scores. Among women, however, GrimAge and DunedinPACE clocks were positively associated with depressive symptoms. Marital status change had a significant negative impact on men’s mental health, leading to about a one-point increase in CESD scores, whereas no significant association was found for women. Furthermore, although the full models did not show significant main effects of marital status in 2016 on CESD scores in 2020, marital history appeared to matter for men. Specifically, men who were separated or divorced in 2016 reported significantly higher CESD scores in 2020 compared to married men, and widowhood also linked to elevated symptoms. These results align with previous findings suggesting that marital dissolution may have more severe mental health consequences for men than for women.

To test Hypothesis 2, which posits that epigenetic clocks moderate the effect of marital status change on mental health, interaction terms between each clock and marital status change were added in models shown in Tables S4 and S5. In the full sample, only the model using Horvath 2 showed a significant moderating effect, attenuating the association between marital status change on CESD scores. However, in the male-only analyses (S4 Table), significant interaction effects were found for Horvath 1, Hannum, Horvath 2, Lin, Garagnani, and GrimAge clocks. Although no significant moderating effects were observed among women, the results suggest that biological aging may buffer the negative mental health impact of marital status change, particularly among men.

Fig 1 graphically illustrates these findings for the male sample. The difference in CESD scores between those who experienced marital status change (solid line) and those who did not (dashed line) was larger among biologically younger men compared to biologically older men. The gap narrowed and reversed when biological age reached approximately 80 years. Before age 80, men who experienced marital status change reported worse mental health than those who did not, whereas beyond age 80, little difference or even slightly better mental health was observed among those who experienced a change. These findings suggest that marital status change is more detrimental to biologically younger men, supporting the moderating role of epigenetic aging.

**Fig 1.**
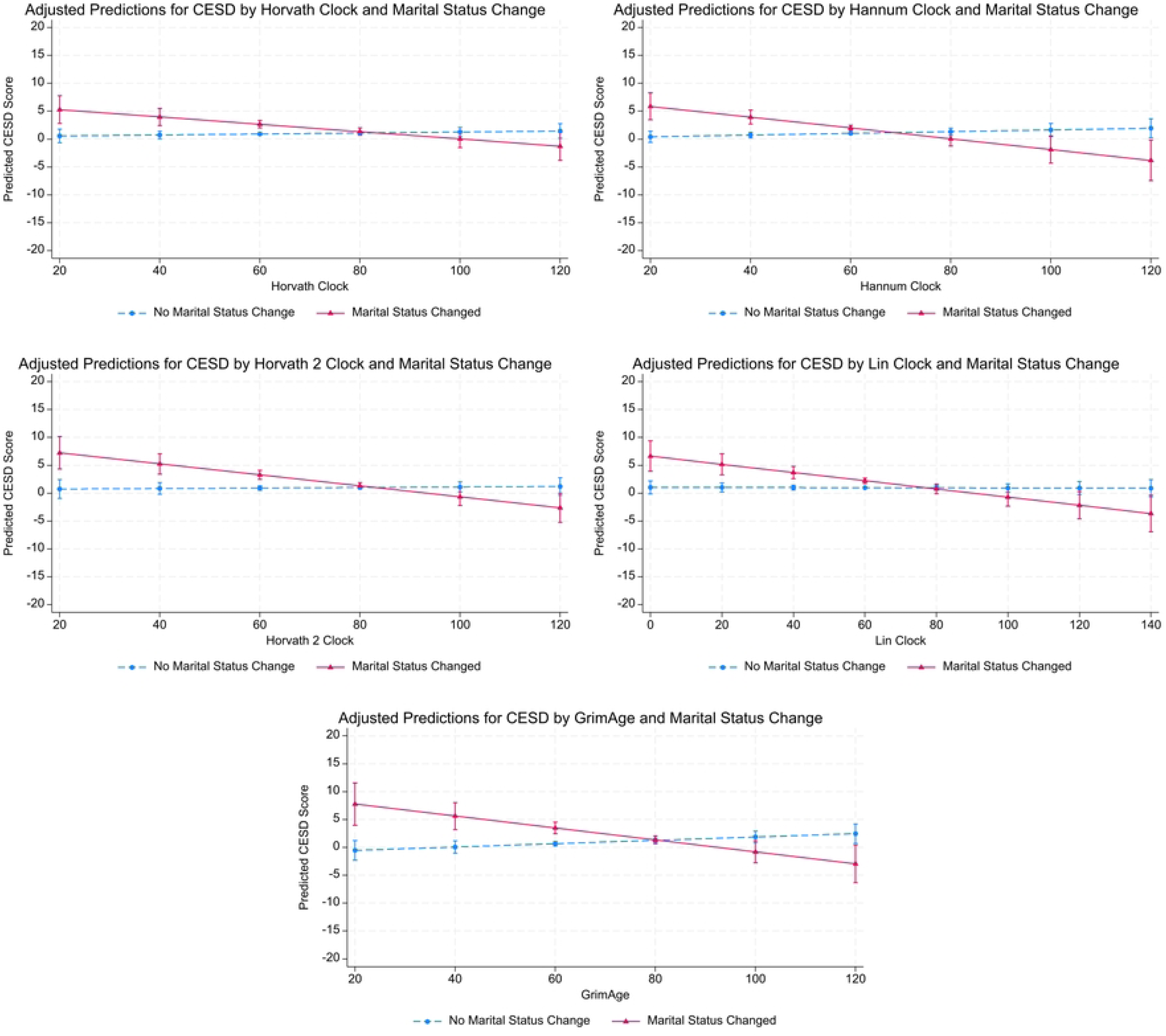
Adjusted Predictions for CESD Score in 2020 by Epigenetic Clocks (2016) and Marital Status Change Between 2016 and 2020 (Males)

### Epigenetic aging, marital status change, and mortality risk

Cox proportional hazards models estimating mortality risk are reported in Tables S6 through S8. As expected, higher epigenetic ages were associated with increased mortality risk after adjusting for covariates. Significant positive associations were observed for the Hannum, Levine, Vidal-Bralo, Zhang, and GrimAge clocks. For instance, a one-year increase in the Levine clock was associated with a 4.3% increase in the hazard of death [exp(0.042) = 1.043]. Similar patterns were found in both male and female models reported in S8 Table.

In contrast to the strong associations observed for epigenetic clocks, marital status change was not significantly associated with mortality risk in the full models, although the coefficients were positive. Thus, while marital transitions may impact mental health, their direct relationship with mortality appeared less pronounced once biological aging was considered.

S7 Table examines the moderating effects of epigenetic aging on the relationship between marital status change and mortality risk to test Hypothesis 2. Significant interaction effects were found for the Horvath 1, Hannum, Horvath 2, Lin, Garagnani, and GrimAge clocks, suggesting that accelerated biological aging may attenuate the association between marital status change on mortality risk. In male-only models (S7 Table), significant moderating patterns were found in seven of the thirteen epigenetic clocks. Even in models where the interaction terms were not statistically significant, the direction of the coefficients was consistent with the hypothesized moderating effect. Among women, no consistent moderating effects were found, except for an interaction using the Zhang clock, which was in the opposite direction.

Fig 2 graphically presents the interaction pattern for the male sample. Survival functions were plotted for men with biological ages of 65 and 85 years, comparing those who experienced marital status change to those who did not. At a biological age of 65, men who experienced a marital transition had lower survival probabilities than those who did not. However, at a biological age of 85, men who experienced marital status change had similar or even better survival probabilities compared to those without a marital transition. This pattern was consistent across models using Horvath 1, Hannum, and Lin clocks. Overall, these results support Hypothesis 2, demonstrating a potential moderating effect of biological aging on the consequences of marital status change for mortality risk. The findings also support Hypothesis 3, as the moderating effects were observed primarily among men, suggesting that marital status transitions may impact men more severely than women and that accumulated life experiences may provide greater resilience for men.

**Fig 2.**
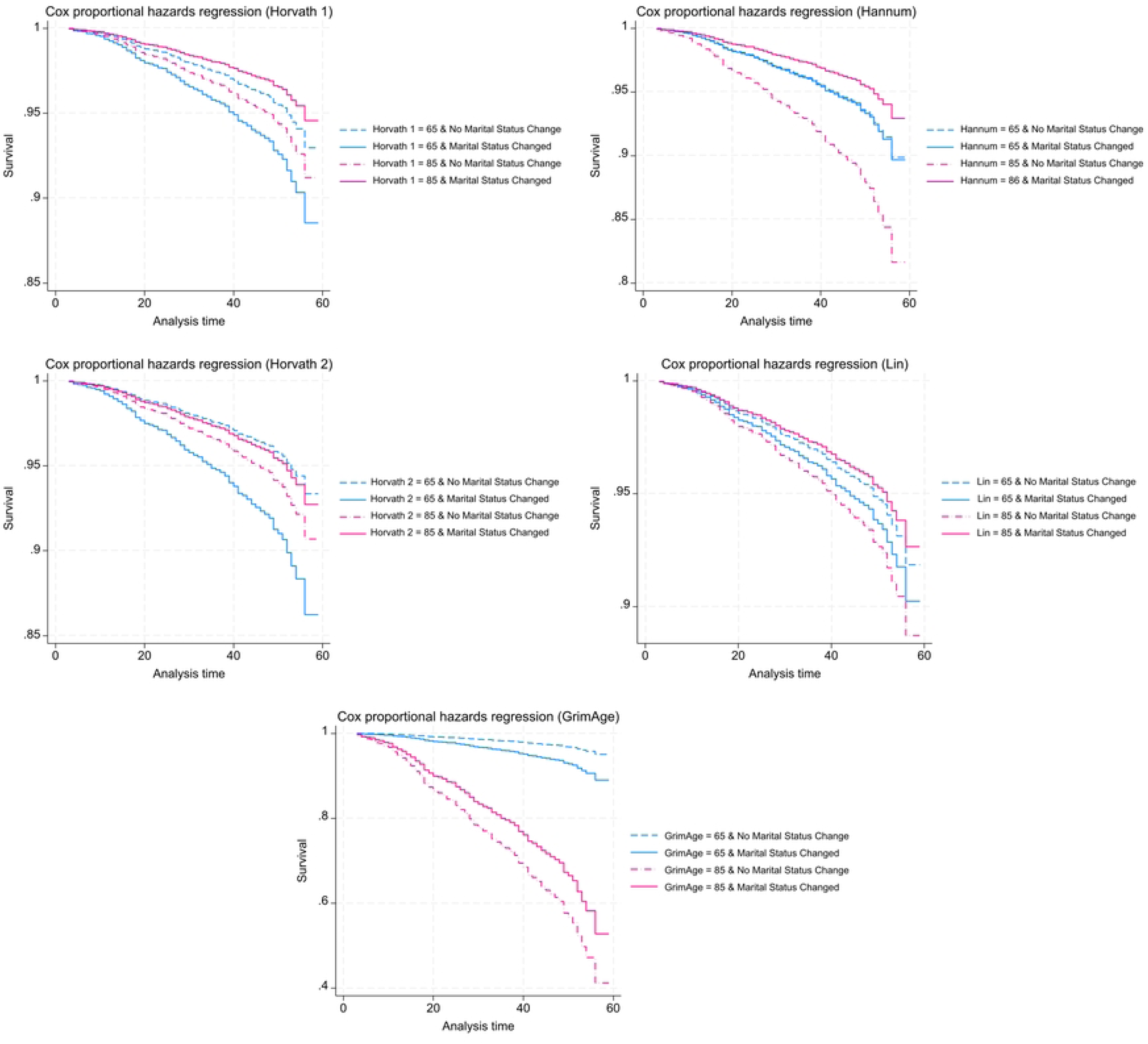
Survivor Function for Mortality Risk by 2020 Interview from Cox Regression by Epigenetic Clocks and Marital Status Change (Males)

### Robustness checks

To assess the robustness of our findings, we conducted several additional analyses (see Supplementary Tables and Supplementary Excel Tables). First, because thirteen epigenetic clocks were tested across multiple outcomes, we applied Bonferroni correction to account for multiple comparisons. Both the main effects and interaction terms were re-evaluated using adjusted significance thresholds. As shown in the supplementary tables, key associations, particularly interaction effects involving GrimAge clock, remained statistically significant after correction, especially among men.

Second, because the analytic sample is relatively small (approximately 1,500 respondents), we used bootstrap resampling with 1,000 replications to assess the stability of the standard errors. Smaller samples can yield less precise estimates and may inflate the risk of Type I or Type II errors. By resampling with replacement, we generated empirical confidence intervals for each coefficient. The direction and magnitude of the main and interaction effects remained consistent across bootstrap samples, suggesting that the findings are not overly sensitive to sample-specific variation.

Third, we conducted two sensitivity checks to address potential survival bias. First, we re-estimated all models using a subsample restricted to respondents younger than 78 years at baseline, corresponding to the U.S. life expectancy in 2016. Although the interaction terms in the Cox models were not statistically significant, their direction remained consistent with the main analyses. The other results also aligned with the primary findings, suggesting that the results were not driven by the oldest respondents. Second, we applied inverse probability weighting to the full sample by modeling the probability of surviving to the 2016 data collection wave using demographic, health, and socioeconomic variables from earlier waves. These weights were incorporated into the CESD and mortality models. The key patterns, including the moderating role of epigenetic clocks, remained robust.

## Discussion and Conclusions

Age may not be as neutral a concept as it initially appears. This study demonstrates that the socially constructed process of aging is embedded within biological systems. By examining the social underpinnings of biological aging and considering epigenetic clocks as markers of accumulated life experiences, four primary findings emerge. First, being separated, divorced, or widowed was associated with significantly elevated epigenetic ages compared to being married, even after adjusting for genetic predispositions and other social factors. Second, marital status change within the previous four years significantly increased depressive symptoms but was not significantly associated with mortality risk. Of the thirteen epigenetic clocks, only GrimAge and DunedinPACE were significantly associated with higher CESD scores, while six were significantly associated with increased mortality risk. Third, although the main effects of epigenetic clocks on depressive symptoms and mortality risk were modest, epigenetic clocks significantly moderated the negative impacts of marital status change, with biologically older individuals coping better with these transitions. Fourth, this buffering pattern was found primarily among men, suggesting potential gender differences in the aging process and the personal meaning of marital transitions.

Consistent with previous research (13–15,28,29,49,50), our study underscores the malleability of epigenetic aging. Even after controlling for chronological age and genetic predispositions, social factors such as marital status and health behaviors were associated with biological aging. Although significant associations were observed for fewer than half of the epigenetic clocks analyzed, the findings suggest that epigenetic aging reflects more than simple biological deterioration. Rather, epigenetic clocks may serve as integrated markers of biological, social, and psychological aging, capturing cumulative life experiences that could include adaptive responses to adversity.

Beyond using epigenetic clocks as biomarkers of physiological deterioration, this study proposes that they also function as indicators of life experience accumulation. Although individuals with older epigenetic ages exhibited higher depressive symptoms and greater mortality risk overall, they also demonstrated greater resilience in the face of marital status changes. This finding resonates with the adage “what does not kill us makes us stronger,” suggesting that accumulated experiences may enhance resilience in later life. The observed patterns persisted even after adjusting for earlier CESD scores and mortality expectations. These results suggest that while aging brings vulnerabilities, it also brings strengths, including accumulated knowledge, coping skills, and emotional regulation. Viewing aging as a resource accumulation process offers a novel perspective on resilience in later life.

This interpretation is supported by theoretical frameworks on resilience, which emphasize the interplay of psychological, social, and biological factors in adaptive functioning (8–10). Resilience develops not only from personal characteristics such as temperament or emotional stability but also from life experiences that foster coping strategies and emotional regulation. In addition to these psychological resources, researchers increasingly recognize a biological dimension of resilience, seen in the body’s capacity to adapt to stress at the physiological level. Findings from recent research (30–32) indicate that resilience-related traits such as self-control and emotion regulation can buffer the negative impact of cumulative stress on epigenetic aging. In this light, the epigenetic moderation effects found in this study further validate the idea that resilience is biologically embedded, with accumulated life experiences strengthening the body’s capacity to withstand future adversities.

Analyses using gender-stratified samples revealed important gender differences in responses to marital status change and in the moderating effects of epigenetic clocks. Overall, marital status change and the interaction between epigenetic clocks and marital transitions were more significant in men’s models than in women’s. These findings are consistent with prior research showing that men may suffer more immediate emotional consequences following marital dissolution (40,51). Specifically, widowed men exhibited older Levine and Vidal-Bralo clocks compared to married men, and marital status change was associated with elevated CESD scores among men but not women.

However, the observation that older biological age buffered the impact of marital transitions for men suggests that, over time, men appear to recover more fully, particularly when accumulated life experiences provide critical resilience resources. This finding aligns with results from (52), who showed that although men experienced greater immediate declines in well-being following divorce, they tended to recover faster than women. In contrast, women experienced prolonged financial setbacks that were slower to resolve. Thus, while men may be more vulnerable in the short term, they appear to rebound more fully when sufficient psychological, social, and biological resources are available.

As a final note, this study is observational in nature. While the analyses identify statistical associations, the results should not be interpreted as evidence of causal relationships.

Although this study offers important insights, it is not without limitations. First, the precise nature of what epigenetic clocks measure remains uncertain. While they can be conceptualized as summaries of cumulative life experiences, the threshold at which accumulated experience becomes detrimental rather than protective remains unclear. Furthermore, given that clocks are based on DNA methylation derived from blood samples, it is unclear how well they reflect aging processes in other tissues or biological systems. Greater understanding of the underlying mechanisms of epigenetic aging is needed.

Second, although we controlled for polygenic scores related to longevity and mental health, it did not control directly for polygenic scores specific to epigenetic clocks themselves, which were unavailable. Future research incorporating these genetic components will be necessary to better isolate the contributions of environmental and social factors on biological aging.

Third, the analytic sample was restricted to individuals of European ancestry, given that the epigenetic clocks and polygenic scores used were primarily developed in European-descent populations. The exclusion of minority groups limits generalizability, and future studies should strive to include more diverse populations, particularly considering the intersectionality of race, ethnicity, and societal inequalities.

Finally, although we sought to integrate biological and social perspectives on aging, further theoretical development is needed to fully conceptualize biological age within a socially constructed framework. Future work should explore how biological and social aging processes interact across the life course, particularly how resilience is shaped by cumulative exposures and resources.

Despite these limitations, this study advances understanding by integrating epigenetic, genetic, and social measures. It demonstrates that epigenetic clocks can be conceptualized as markers of accumulated life experiences and highlights how these experiences may function as resilience resources in later life. Recognizing aging as both a biological and social process, and as a potential source of strength rather than inevitable decline, represents an important step forward in aging research.

## Data Availability

The Health and Retirement Study data used in this study are available to the public at the following link: https://hrs.isr.umich.edu/data-products.

## Notes

* Declarations of interest: none.

**Funding Statement:** This research was supported by a grant from the National Science and Technology Council of Taiwan, grant number 113-2628-H-002-016-.

**Conflict of Interest:** None.

### Competing Interest Statement

The authors have declared no competing interest.

### Author Declarations

This study was reviewed and approved by the Research Ethics Committee of National Taiwan University (NTU-REC No.: 202401HS013) and was classified as exempt on January 17, 2024. A waiver of informed consent was granted.

